# Changes in eating habits and lifestyles in Peruvian population during social isolation by the COVID-19 pandemic

**DOI:** 10.1101/2021.03.08.21252979

**Authors:** Salomón Huancahuire-Vega, Edda E. Newball-Noriega, Ricardo Rojas-Humpire, Jacksaint Saintila, Mery Rodriguez Vásquez, Percy. G. Ruiz-Mamani, Wilter C. Morales-García, Michael White

## Abstract

**Background:** The COVID-19 pandemic caused that some governments have implemented house confinement measures with probable consequences on lifestyle, particularly affecting eating habits, physical activity, sleep quality, and mental health.

**Objectives:** The aim of this study was to assess the frequency of lifestyles, physical activity and sleep characteristics, as well as changes in eating habits in the Peruvian population during COVID-19 pandemic.

**Methods:** A Cross-sectional descriptive study was performed. We analyzed adults from Peru between July to August 2020 based on an online self-administered questionnaire divided into sociodemographic, anthropometrics and COVID-19 diagnostic reported, lifestyle habits and frequency of consumption of foods.

**Findings:** During confinement by COVID-19, 1176 participants were studied, 39% were student, 37.5% were workers and 46% were assert not to work. The population asserted gain weight (1 to 3 Kg) and 35.7% were overweight. The lifestyles habits showed that 54.8% affirmed to doing physical activity and a large proportion (37.2%) asserted sleep less. The Peruvian population presented a main feeding patter of breakfast (95.7%), lunch (97.5%), dinner (89.1%) and brunch (44.9%). Likewise, feeding habits before and during COVID-19 pandemic showed that vegetables (OR:1.56, CI95% 1.21 - 200), fruit (OR: 1.42, CI95% 1.10 – 1.81), legumes (OR:1.67, CI95% 1.23 – 2.28) and eggs (OR: 2.00, CI95% 1.52 – 2.65) presented significantly consumption increase during social isolation, while bakery products (OR: 0.74, CI95% 0.56 – 0.97), meat, snack, refreshment and fast-food decrease consumption. Other food no significant differences were presented.

**Conclusion:** This study in a Peruvian population showed an important frequency of overweight and sleep disorders. There was a slight increase in physical activity despite the social isolation measures and an increase in health eating habits, nevertheless a majority reported gaining weight.

## 1. Introduction

An announcement made by the WHO on January 30, 2020 declared COVID-19 a global health emergency due to the exponential growth of cases in China and other countries of the world.^1^ As of October 18, 2020, according to the Coronavirus Resource Center at Johns Hopkins University, more than 39 million cases have been confirmed, while the number of deaths has risen to more than 1 million.^2^ Given the circumstances measures needed to control the spread of the disease, government authorities were forced to take decisive actions such as social distancing and mandatory isolation in homes, which have led to restrictions on various daily activities, with probable consequences on lifestyle, particularly affecting eating habits, physical activity, sleep quality, and mental health.^3^

In Peru, the first case of COVID-19 infection was reported by the government on March 6, 2020. Since then, the government has implemented a series of preventive measures in order to limit and contain the spread of the disease. On March 16 the government, through Supreme Decree No. 008-2020-SA, declared a nationwide state of health emergency for a period of ninety (90) calendar days, dictating prevention measures and mandatory social isolation (national quarantine) for the control of COVID-19.^4^ As of June 26, the government announced an extension that modified the restriction to a targeted social isolation, but the state of health emergency and social distancing measures were extended until December 31.^5^

The period of social isolation due to COVID-19 has caused a series of changes in people’s daily routine and lifestyle.^6^ Available evidence has shown that during the pandemic, physical activity decreased in the population, while the amount of screen time increased significantly.^6^ In fact, prior to the pandemic, these factors, along with obesity, were described as a public health problem in the Peruvian population, and COVID-19 may make this situation even worse.^7^ In addition, studies have shown the negative impact of sedentary behaviors on physical and mental health.^8^ Likewise, the fear caused by COVID-19 has an indirect effect, causing negative emotions by affecting the quality of sleep, contributing to an increase in obesity.^9^

Additionally, evidence has shown that the risks of choosing inappropriate eating behaviors can increase as a psychological and emotional response to COVID-19.^10^ In fact, it has already been shown that negative emotional experiences can lead to overeating.^11^ Eating habits characterized by a high consumption of ultra-processed, high-calorie-density foods and low intake of fiber, antioxidants and other bioactive elements can affect the immune system, causing chronic activation of the innate system and inhibition of the adaptive immune system response by increasing oxidative stress, thus delaying defensive intervention against pathogens.^12^

Previous studies have demonstrated the beneficial effects of physical activity and sleep quality on mental health.^13^ In addition, adequate physical activity can improve the quality of sleep, reducing the risk of morbidity from chronic non-communicable diseases.^14^ Furthermore, appropriate eating habits can be a protective factor for physical and mental health, disease prevention and strengthening of the immune system.^15^ Indeed, improved lifestyle, as a possible strategy that includes regular physical activity, less screen time, adequate rest and proper dietary habits through a diet based on consumption of minimally processed plant foods and healthy fats, would lead to better control of chronic noncommunicable diseases and thus strengthen the immune system during the COVID-19 pandemic.^15^ The objective of the present study was to evaluate the frequency of lifestyles, physical activity and sleep characteristics, as well as changes in eating habits in the Peruvian population during the period of social isolation due to the COVID-19 pandemic.

## 2. Materials and Methods

### 2.1. Design and participants

This is a cross-sectional descriptive study based on a self-administered questionnaire. This questionnaire was addressed to the Peruvian adult population (over 18 years of age), and was applied using a non-probabilistic convenience sampling. Responses from 1218 participants were obtained, of which 42 were eliminated due to incompleteness and certain inconsistencies related to age, height, weight, etc.

The study followed the international ethical recommendations contained in The Declaration of Helsinki. All participants were informed about the characteristics of the study as well as their anonymous, voluntary and confidential participation. This study was approved by the Ethics in Research Committee of the Universidad Peruana Unión (No. 2020-CEUPeU-00013).

### 2.2. Procedure

The questionnaire was applied from July 16 to August 31, 2020 (period in which the Peruvian population was in partial confinement) using the Google form platform and disseminated through social networks (Facebook and Whastapp) and institutional mailing lists. The questionnaire was completed through any electronic device with an internet connection. On the initial page, the participants were explained the voluntary nature of their participation with informed consent, as well as the justification, objectives, possible risks and benefits of the study before continuing with the instrument itself.

### 2.3. Questionnaire and variables

The questionnaire was developed by the authors to specifically address the particularities of the current pandemic context and its impact on lifestyle. The questionnaire included 42 questions divided into four sections: Sociodemographic data (10 questions: gender, marital status, age, level of education, occupational status, employment status, if the social isolation was carried out, duration of social isolation, people in the home before and during social isolation); anthropometric data and COVID-19 diagnosis reported (5 questions: weight, height, weight change during social isolation, number of kilograms, COVID-19 diagnosis); information on lifestyle habits (11 questions: physical activity, change and hours of sleep, meals eaten per day during confinement); information on the frequency of consumption of the main food groups (16 questions: vegetables, fruits, legumes, dried fruits/nuts, meat, fish, eggs, milk, yogurt, bakery products, snacks, beverages, beer, wine, other alcohol, fast food) before and during social isolation, based on five response categories: Participants who reported having consumed food from 1 to 4 times/week were considered as low consumption, while high consumption was considered when they reported having consumed from 5 times/week to 2 or more times/day.^16^

### 2.4. Statistical analysis

The data analysis was performed in RStudio v4.0.2 (R Foundation for Statistical Computing, Vienna, Austria; http://www.R-project.org). The variables were encoded and analyzed such as categorical. To descriptive analysis categorical variables were cluster in to tables with absolute and relative frequency (%). To comparative analysis χ^2^ or Fisher’s exact test were used to categorical variables depending of distribution of variable. Subsequently, to determinate differences before and during COVID-19 pandemic McNemar test was performed. A p<0.05 was considered statistically significant.

## 3. Results

A summary of the sample’s sociodemographic characteristics can be found in Table 1. In total 1176 participants were studied, 571 men and 605 women. The largest age groups were 18 to 25 years old (44.4%) with predominantly women and 26 to 40 years old (33.7%) with predominantly men. Overall, 67.7% were single and 38% had completed university studies. During social isolation, 39% of those sampled were students, 37.5% were workers, 30.8% worked with a formal contract, 46% were assert not to work and 67.6% were under social isolation for more than 61 days as of the date of the study.

**Table 1.**
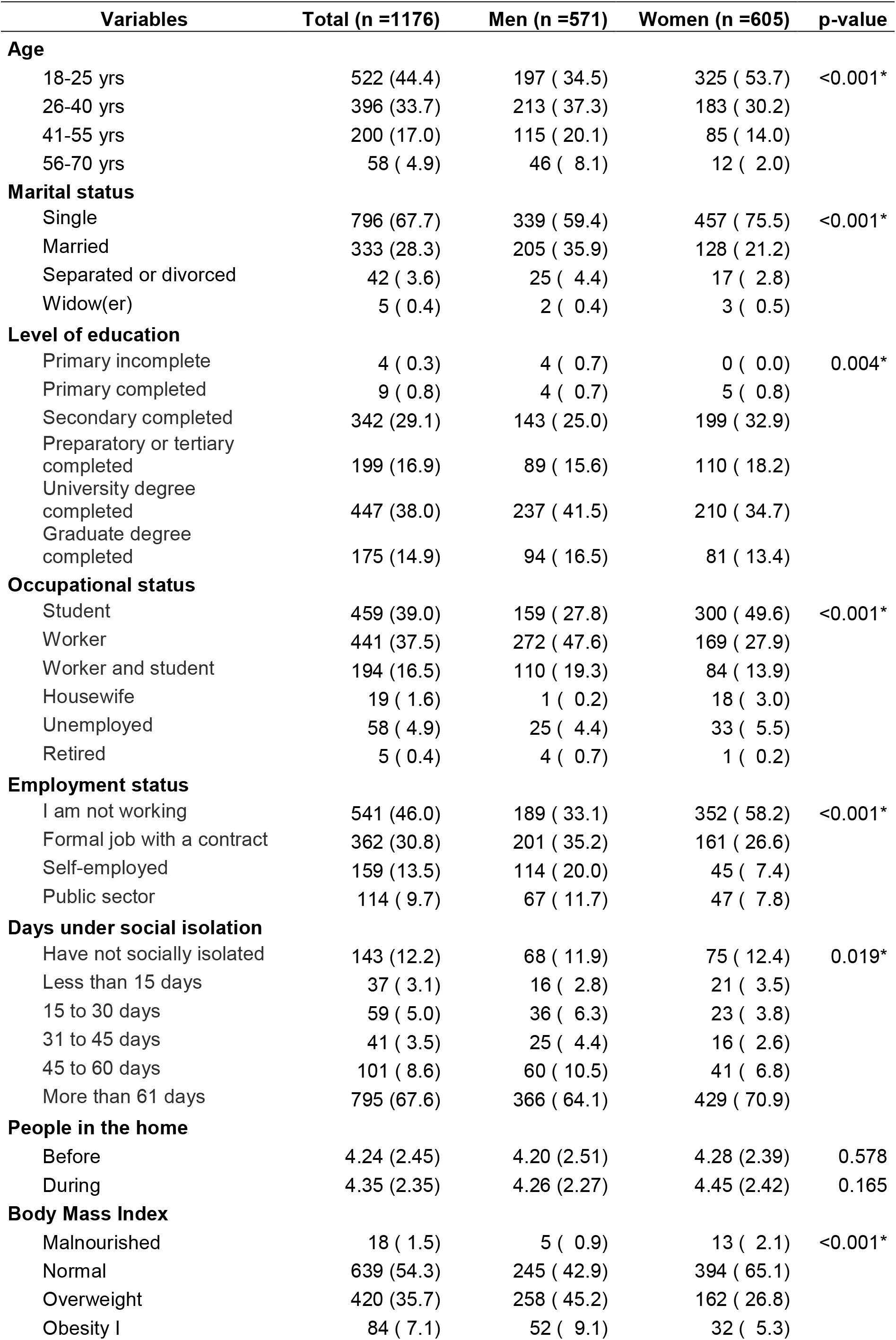
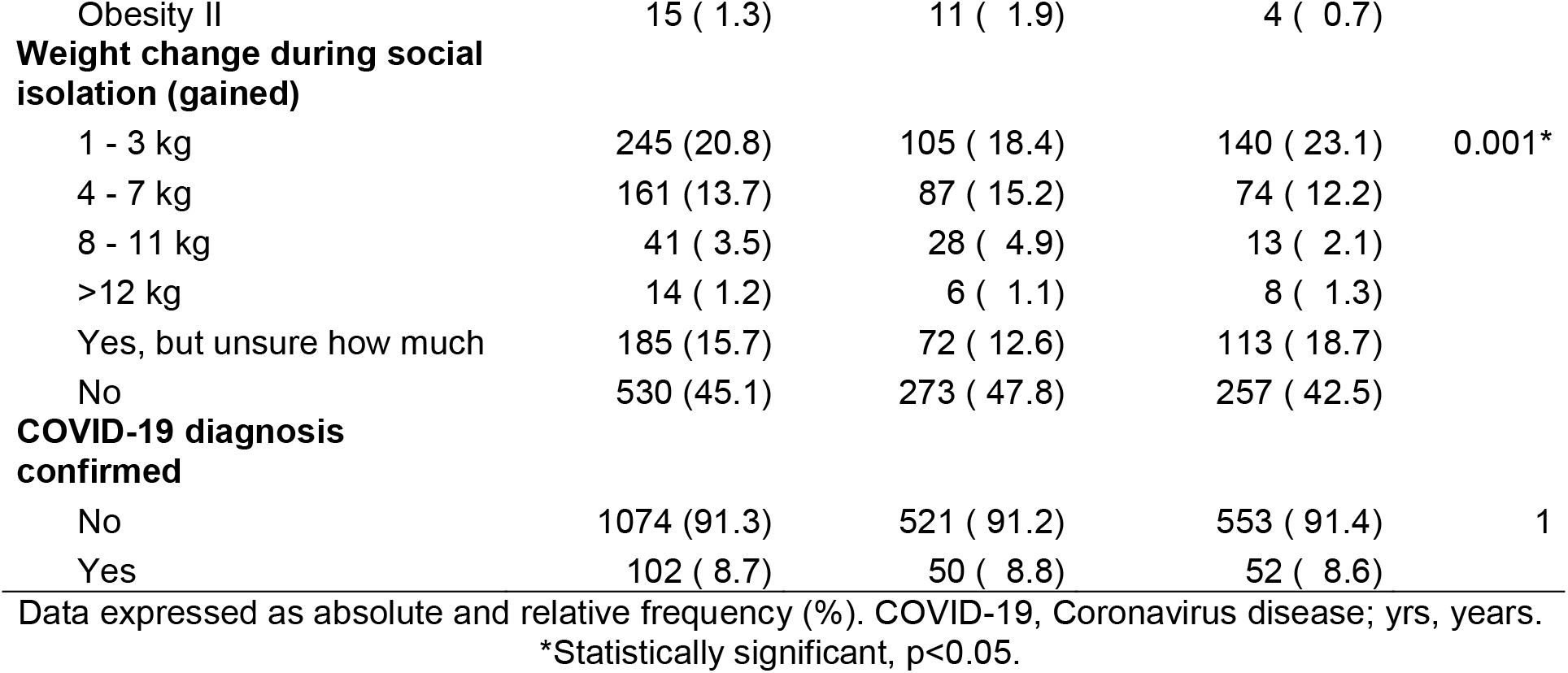
Sociodemographic and anthropometric data.

A total of 35.7% of those studied were overweight and 8.4 % were obese, with a majority of men in both groups. A large portion reported having gained weight, 20.8% and 13.7% between 1 to 3 kg and 4 to 7 kg respectively. 8.7% of those surveyed had a confirmed diagnosis of COVID-19.

Some aspects of lifestyles showed patterns and changes when grouped by sex during the social isolation (Table 2). More than half of those studied affirmed to doing physical activity (54.8%) between 1 to 2 (14.7%) and 3 to 4 (26.4%) days per week, for at least 30 min. The reported sleep routine presented significate changes wherein 37.2% responded that they slept less and 36.9% slept more hours at night compared to before the pandemic. The hours of sleep per night were largely between 4 to 6 (35.6%) and 7 to 9 (55.4%) hours.

**Table 2.** Lifestyles during social isolation.

| Variables | Total (n =1176) | Men (n =571) | Women (n =605) | p-value |
| --- | --- | --- | --- | --- |
| **Physical activity** |  |  |  |  |
| No | 532 (45.2) | 258 ( 45.2) | 274 ( 45.3) | 1 |
| Yes | 644 (54.8) | 313 ( 54.8) | 331 ( 54.7) |  |
| **Days per week** |  |  |  |  |
| None | 532 (45.2) | 258 ( 45.2) | 274 ( 45.3) | 0.204 |
| 1 to 2 days | 173 (14.7) | 76 ( 13.3) | 97 ( 16.0) |  |
| 3 to 4 days | 310 (26.4) | 147 ( 25.7) | 163 ( 26.9) |  |
| 5 to 6 days | 131 (11.1) | 71 ( 12.4) | 60 ( 9.9) |  |
| 7 days | 30 ( 2.6) | 19 ( 3.3) | 11 ( 1.8) |  |
| **Duration** |  |  |  |  |
| No physical activity | 532 (45.2) | 258 ( 45.2) | 274 ( 45.3) | 0.425 |
| Less than 30 minutes | 257 (21.9) | 122 ( 21.4) | 135 ( 22.3) |  |
| 30 minutes to 1 hour | 335 (28.5) | 159 ( 27.8) | 176 ( 29.1) |  |
| 1 to 2 hours | 46 ( 3.9) | 28 ( 4.9) | 18 ( 3.0) |  |
| Other | 6 ( 0.5) | 4 ( 0.7) | 2 ( 0.3) |  |
| **Sleep routine** |  |  |  |  |
| No change | 304 (25.9) | 171 ( 29.9) | 133 ( 22.0) | 0.006* |
| Sleep more | 434 (36.9) | 204 ( 35.7) | 230 ( 38.0) |  |
| Sleep less | 438 (37.2) | 196 ( 34.3) | 242 ( 40.0) |  |
| **Hours of sleep** |  |  |  |  |
| 0 to 3 hours | 25 ( 2.1) | 12 ( 2.1) | 13 ( 2.1) | 0.170 |
| 4 to 6 hours | 419 (35.6) | 188 ( 32.9) | 231 ( 38.2) |  |
| 7 to 9 hours | 651 (55.4) | 335 ( 58.7) | 316 ( 52.2) |  |
| >10 hours | 81 ( 6.9) | 36 ( 6.3) | 45 ( 7.4) |  |

Data expressed as absolute and relative frequency (%). \*Statistically significant, $p < 0.05$

Eating patterns during the social isolation showed that approximately 90 to 95% ate breakfast, lunch and dinner. Additionally, some extra food consumption was reported for brunch (42.9 to 46.8%), afternoon meal (38.2 to 39.8%) and snacks (35.2 to 40.8%), these results presented no differences by sex (Figure 1).

**Figure 1.**
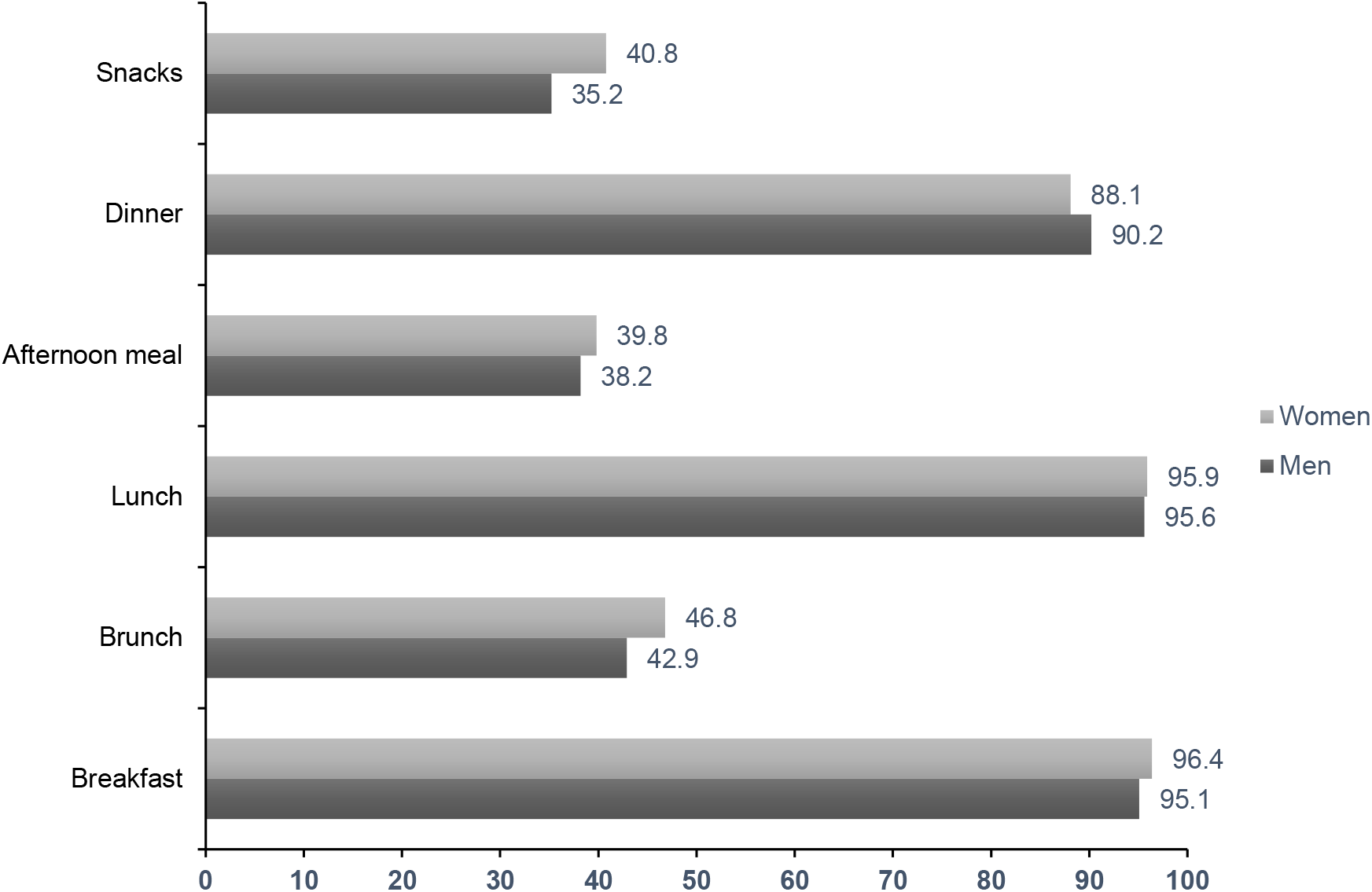
Frequency of eating patterns during social isolation.

The eating habits showed some changes before and during social isolation (Table 3). Fruit and vegetables presented elevated frequency of consumption; while, legumes and dried fruits/nuts showed low frequency of consumption (Figure 2). On the other hand, just vegetables (OR:1.56, CI95% 1.21 - 200), fruit (OR: 1.42, CI95% 1.10 – 1.81) and legumes (OR:1.67, CI95% 1.23 – 2.28) presented significantly consumption increase during social isolation. Approximately a quarter of the population presented high consumption of meat and eggs (Figure 3); while, dairy products and fish showed low frequency of consumption. In the animal-based foods only eggs (OR: 2.00, CI95% 1.52 – 2.65) presented significantly increasing during social isolation. The consumption of alcoholic drinks and ultra-processed food were the lowest; while, approximately 20% and 40% of population presented high consumption of beverages and bakery products, respectively (Figure 4). In the processed food, just bakery products (OR: 0.74, CI95% 0.56 – 0.97) presented change before and during social isolation.

**Figure 2.**
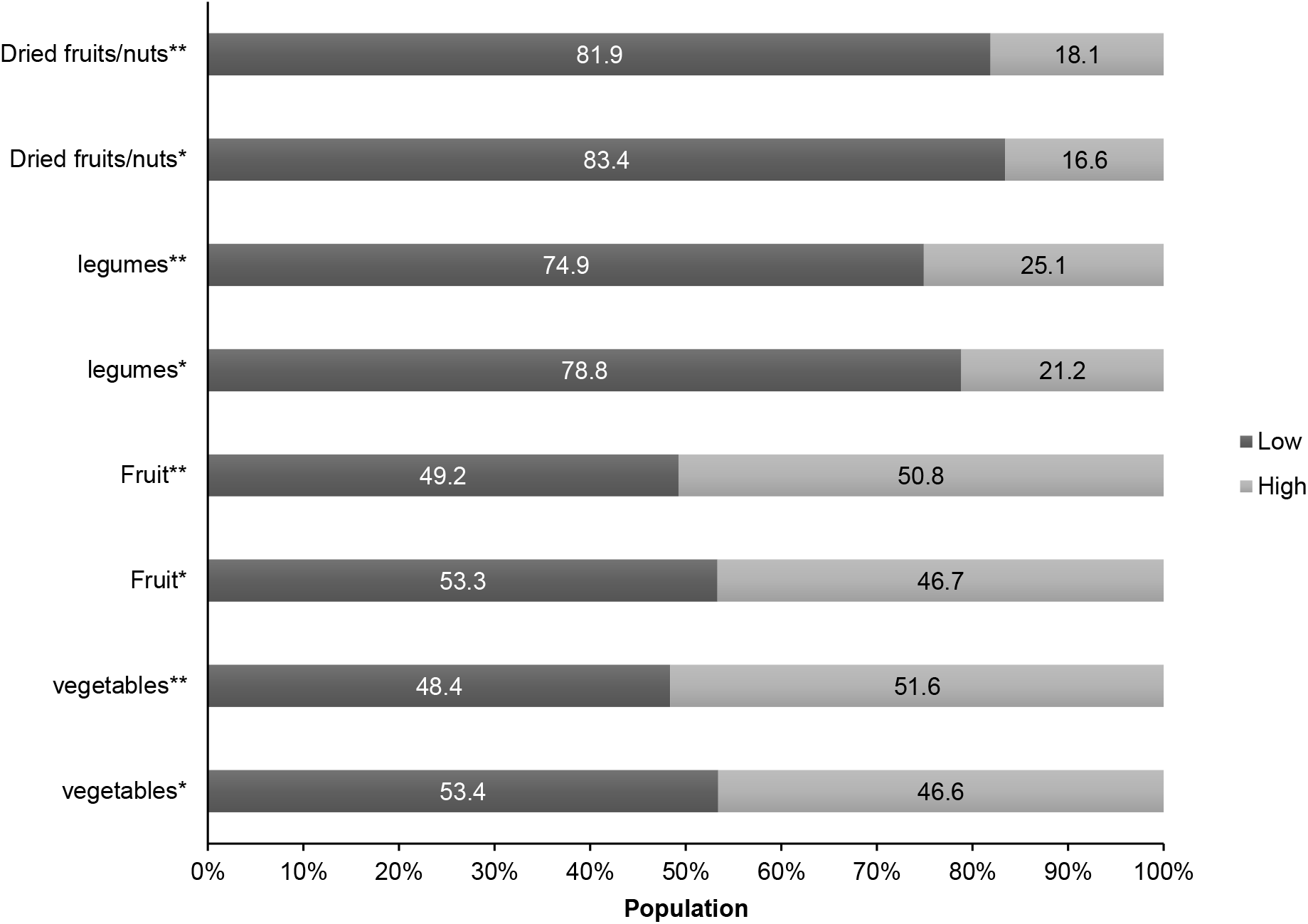
Changes in the frequency of low and high consumption of plant-based foods before and during social isolation. *Before social isolation, **During social isolation.

## 4. Discussion

The objective of this study was to evaluate the frequency of lifestyles, physical activity and sleep characteristics, as well as changes in eating habits before and during social isolation due to the COVID-19 pandemic in a Peruvian population.

The findings of this study show that more than 60% of those surveyed remained at home, as a result of the imposition of social isolation due to outbreaks of COVID-19, and thus despite the more than 61 days since the start of the pandemic in Peru, only 8.7% had a confirmed diagnosis of COVID-19. Studies indicate that sustained transmission in the early stage of the epidemic in Peru (March 2020) showed an exponential growth dynamic despite non-pharmaceutical interventions by the authorities such as compulsory social isolation.^17^ In addition, Peru had some of the highest infection and mortality rates in the world, along with countries such as Italy, Spain, Brazil, the United Kingdom and France.^18^

This study highlights the potential impact of social isolation due to COVID-19 by including a variety of weight-related behaviors in Peruvian adults. The results of this study indicate that of the total number of participants subjected to social distancing, 35.7% were overweight and 8.4% were obese. Likewise, the findings show that the imposition of social isolation influenced lifestyles causing negative effects and weight changes, the population reported gaining weight with 20.8% between 1 to 3 kg and 13.7% from 4 to 7 kg respectively. Similar studies in 765 patients in Kurdistan, Iraq indicated a weight gain during social isolation of less than 2 kg in almost all patients (98.05%) and those who gained more than 3 kg were mostly women or from the center of large cities.^19^ Another study on the BMI of a Lebanese population showed 30.8% overweight and 12.9% obese.^20^ On the other hand, 32% of the sample population (2364 participants) in a study in the United Kingdom was classified as obese during social isolation.^21^ In addition, a perceived increase in body weight was reported in the United States with an average body mass index (BMI) of 25.8.^22^

Weight gain is associated with BMI, as those with a higher BMI gain weight more often compared to those with a normal BMI. Studies prior to the pandemic showed a prevalence of overweight and obesity in Peruvians ≥ 20 years old and it was women who had a higher prevalence of obesity and abdominal obesity than men.^23^ The results of this study indicate a weight gain of between 1 and 7 kg, it is likely that high rates of stress and depression will push people to consume foods with high sugar content and generate lifestyle changes and negative psychological impacts (increased prolonged naps, fears of infection, frustration, boredom, inadequate information, financial losses, negative psychological effects, etc.), causing long-term effects on obesity and the presence of co-morbidities, making obesity a higher risk of death related to the COVID-19 pandemic.^24^

The association of severe COVID-19 cases with higher than normal BMI values has been demonstrated.^25^ For example, 88.2% of patients admitted to Wuhan Union Hospital between January and February 2020 who were not survivors of this illness had a BMI > 25 kg/m^2^. ^26^ Some explanations state that a higher than normal BMI increases susceptibility to respiratory infections due to the deterioration of lung mechanics and higher concentrations of pro-inflammatory molecules.^27^ On the other hand, the excessive amount of adipose tissue in the abdominal area decreases diaphragmatic excursion and affects the ventilation of the base of the lung, which can lead to hypoxemia, clinical characteristics observed in this illness.^28^

Pre-pandemic studies showed that the Peruvian population had a high prevalence of physical inactivity and sedentary behaviors.^29^ This study found substantial changes in behavior during social isolation, the majority of participants claimed to be physically active (54.8%) for 1-2 (14.7%) or 3-4 (26.4%) days per week, for at least 30 minutes. Similar studies showed increased regular participation in moderate and vigorous physical activity in older adults during quarantine.^30^ On the other hand, other studies show that total physical activity decreased significantly during social isolation, due to greater sedentary behavior, more prolonged during the weekend.^31^ While some individuals report a decrease in physical activity due to mobility restrictions in the context of COVID-19, others report increases in physical exercise and other efforts to “stay fit”.^32^

In most studies, social isolation induced a reduction in physical activity and an increase in sitting time, or only low-intensity activities such as housework.^31^ This sudden cessation of physical exercise is detrimental in the context of this disease, since this decrease is associated with rapid muscle atrophy and reduced energy consumption by the muscles, promoting obesity and lipid accumulation, decreased ability of organ systems to resist viral infection, increased risk of damage to the immune system, respiratory system, cardiovascular system, musculoskeletal system and brain.^33^ Therefore, the WHO recommends at least 150 minutes per week of moderate to vigorous intensity physical activity or 75 minutes of high-intensity physical activity per week, or a combination of both.^34^

More than 70% of the participants in this study report substantial changes in sleep patterns during the period of isolation due to COVID-19. Similar studies revealed a high prevalence of anxiety sleep disorders and depressive symptoms in the population during the COVID-19 social isolation period, with women reporting a higher risk of sleep difficulties,^35^ which was also observed in our study. It is likely that women were more receptive to expressing psychological distress and somatic symptoms.^36^ On the other hand, the uncertainty of being infected by the coronavirus would cause greater pressure in the population, relating to hypochondriacal concerns, sleep apnea, and childhood trauma that can be reflected in sleep mentality, affecting its quality.^37^ Sleep plays a critical role in physical and mental health, and adequate sleep duration and quality are essential to cope with major life events such as the COVID-19 pandemic.^38^

This study also showed significant changes in the eating habits that this Peruvian population adopted during social isolation, among which the increase of eggs, legumes, fruit and vegetables, as well as the decrease of bakery products. Some studies observed an improvement in the pattern of consumption of healthy foods and a restriction of unhealthy foods, especially in the younger population (<30 years).^39^ In contrast, studies in Italy indicated changes in eating habits in almost half the population, so that women were more likely than men to increase their food intake to feel better.^8^ In other countries, after-dinner snacks, carbohydrate consumption, consumption of sweets, cookies, and cakes, and consumption of fresh food products increased.^40^ The lack of dietary and eating restriction may be due to a response to stress and reduced physical activity, which could lead to decreased nutritional quality.^40^ This unhealthy behavior is observed in other studies primarily in adult populations with obesity and in settings with a lower eating environment index.^21^

In addition to changes in eating habits, alcoholic drinks and ultra-processed food consumption was the lowest and showed no changes before and during social isolation. Some studies suggest that increased alcohol consumption during the COVID-19 pandemic may be a response to boredom, inactivity, isolation, and as a means of combating anxiety, stress, and/or loss of sleep as a result of containment measures introduced to minimize the spread of the virus.^41^

Healthy lifestyles are relevant to various diseases as well as to maintaining people’s mental health. Among these, eating habits may be a factor contributing to the geographical variation of COVID-19 infection rates, as the consumption of certain foods may be associated with lower mortality. A balanced diet is vital for the prevention of COVID-19, related chronic diseases and has a favorable impact on COVID-19 mortality.^42^

The main limitation of this work is the lack of representativeness of the population sample. Therefore, it is not possible to expand the results to the population of the country. There is also an over-representation of younger individuals (< 25 years). Moreover, all data collected are self-reported and this could make them not completely reliable, especially when it comes to reporting behaviors for which there may be a social stigma (e.g., alcohol and tobacco consumption). Another limitation could be the lack of information about religious practices which could have influenced diet and lifestyle changes.

In conclusion, this study in a Peruvian population showed an important frequency of overweight and sleep disorders. On the other hand, efforts to included physical activity despite the social isolation measures, an increase in health eating habits, but nevertheless a majority reported gaining weight. Future studies should consider additionally potential factors, starting with the observation in this study that most participants had a changed sleep pattern during social isolation, and potentially including other factors like stress, type of exercise, pedometer step counts, and portion sizes.

## Data Availability

The data presented in this study are available on request from the corresponding author

## Author statements

## Funding

This research received no external funding.

## Data Availability Statement

The data presented in this study are available on request from the corresponding author.

## Competing interests

The authors have no conflict of interest

## Notes

### Competing Interest Statement

The authors have declared no competing interest.

### Funding Statement

Nothing to declare

### Author Declarations

The study followed the international ethical recommendations contained in The Declaration of Helsinki. All participants were informed about the characteristics of the study as well as their anonymous, voluntary and confidential participation. . This study was approved by the Ethics in Research Committee of the Universidad Peruana Union (No. 2020-CEUPeU-00013).

